# Mental Health Literacy questionnaire short-version: psychometric properties in young adults from Brazil, Germany, Greece, Lithuania, Malaysia, Nepal, Nigeria, Pakistan, Poland, Serbia and Turkey

**DOI:** 10.1101/2025.02.25.25322614

**Authors:** Diogo Costa, Marisa Costa, Wanderson Carneiro Moreira, Maria do Perpétuo S.S. Nóbrega, Birthe Fritz, Michèle Wessa, Argyroula Kalaitzaki, Flora Meintani, Olamide Aturu, Teresa Sui Mien Yong, Mohammad Zabri Johari, Sandesh Dhakal, Dev Bandhu Poudel, Mujeeba Ashraf, Emilia Soroko, Marina Kovacevic Lepojevic, Ana Radanovic, Nevin Gunaydın, Funda Özpulat, Luísa Campos, Pedro Dias

## Abstract

**Objectives:** The prevalence of mental health disorders has increased in recent years, representing a major public health concern. Increasing the literacy about mental health signs and symptoms is paramount for effective prevention. Having previously studied the psychometric properties of the Mental Health Literacy Questionnaire for young adults – short version (MHLq-SVa) in six countries, we now extend the validity study of this 16-item scale to eleven countries, namely Brazil, Germany, Greece, Lithuania, Malaysia, Nepal, Nigeria, Pakistan, Poland, Serbia and Turkey.

**Study Design:** Multicentre cross-sectional study

**Methods:** Translated and adapted versions of the MHLq-SVa were administered to young adults aged 17-25 years in all countries, totalling 5054 (72% women) with full information for all scale items. Confirmatory factor analysis and internal consistency (Cronbach’s alphas) were computed to test the scales’ four-dimensional structure (Knowledge of mental health problems; Erroneous beliefs/stereotypes; First-aid skills and help-seeking behaviour; Self-help strategies) and reliability.

**Results:** Goodness-of-fit indices for the four-factor solution model suggested good fit in all countries. Exceptions were Pakistan with higher Root mean squared error of approximation and Standardized root mean squared residual, and Brazil with a higher chi-square value and lower Comparative fit index.

**Conclusions:** The construct validity of the MHLq-SVa was supported by data from eleven countries, adding to previous research on the MHLq-SVa psychometrics, suggesting cross-cultural reliability as a Mental Health Literacy assessment tool.

## Introduction

The prevalence and burden of mental health disorders have increased in recent years worldwide, representing a major public health concern. Mental health conditions are now one of the leading causes of disability and contribute substantially to the global burden of disease.^1,2^ Despite their high prevalence, mental health disorders often remain underdiagnosed and undertreated, largely due to several interrelated barriers. The pervasive stigma and discrimination faced by individuals diagnosed with mental health conditions is a significant barrier. A general lack of knowledge about the features of mental illness and limited awareness of how to access appropriate treatment are additional barriers ^3–6^. The relationship between these factors has been suggested to vary cross-culturally ^7^. Therefore, it is imperative to implement strategies to enhance the public’s knowledge, attitudes, and subsequent behaviour toward mental disorders, which in turn affects access to mental healthcare ^4^. These skills are known as mental health literacy (MHL). This multifaceted concept has been proposed to encompass knowledge of mental health issues, recognition of signs and symptoms of mental illness, understanding of treatment and care options, and knowledge of self-help and first-aid strategies to assist individuals experiencing mental health problems ^8^.

Research has extensively examined variables influencing MHL levels, revealing significant patterns. Gender differences have emerged, with young female adults showing higher MHL levels compared to their male counterparts.^9–13^ Proximity to individuals with mental health conditions is another critical factor; individuals who have friends or family members with mental health problems tend to display higher MHL, likely due to increased awareness and empathy.^10,14^ Cultural aspects further influence MHL, with studies indicating that personal beliefs, religious practices, language, and subjective experiences significantly affect how mental health issues are perceived and addressed.^15,16^ The development of valid and reliable tools to measure MHL is essential for identifying knowledge gaps, evaluating interventions, and facilitating cross-cultural comparisons. Despite the existence of other nationally validated scales assessing MHL, there is still a need for cross-country studies to examine the validity and reliability of standardized tools ^17^. Previously, the Mental Health Literacy Questionnaire for Young Adults (MHLq-YA) demonstrated robust psychometric properties.^18^ Subsequently, a short version of this scale (MHLq-SVa) was validated in six countries: China, India, Indonesia, Portugal, Thailand, and the United States, providing a concise yet comprehensive instrument for measuring MHL.^19^ Building on this foundation, the present study aims to expand the validity and applicability of the MHLq-SVa by examining its psychometric properties in eleven additional countries: Brazil, Germany, Greece, Lithuania, Malaysia, Nepal, Nigeria, Pakistan, Poland, Serbia and Turkey.

## Methods

This was a cross-sectional survey. In each country, the 29-item MHLq-YA or the 16-item MHLq-SVa questionnaires were translated from English or Portuguese to the main language, back-translated, and piloted. A semantic comparison of the translated and back-translated versions followed, and the resulting data were then reviewed by experts. The translation process followed previously established standard procedures^20^.

Participant recruitment was conducted online in all countries except Serbia and Turkey, where participants were recruited in person. A convenience and snowball sampling procedure was employed, whereby potential participants were requested to recruit from their contacts and networks. The questionnaire was administrated online (web-form questionnaire, ex: SoSiSurvey platform), and advertised via social media platforms (such as Facebook, WhatsApp and Instagram), news websites, and various webpages in all countries except Turkey. In Germany, in addition to social media platforms, flyers were distributed for recruitment, QR codes were created for students, a database of the Gutenberg Brain Study was accessed, emails were sent to clinics and adverts were posted in specialist journals for healthcare professionals. The data collection period varied across countries, lasting from two (e.g.: Turkey, Pakistan, Serbia) to eight months (e.g.: Brazil).

In addition to the MHLq-YA, participants reported their gender and age.

The short version of the MHLQ-YA includes 16 items^19^, derived from the original 29-item version^18^. The 16-item version retains the assessment of four mental health literacy dimensions, namely:

a. knowledge of mental health problems (e.g., “Mental disorders affect people’s thoughts.”; “Changes in brain function may lead to the onset of mental disorders.”),
b. erroneous beliefs/ stereotypes (e.g., “Mental disorders don’t affect people’s behaviours.”; “Only adults have mental disorders.”,
c. help-seeking and first-aid skills (e.g., “If I had a mental disorder, I would seek a psychologist’s help.”; “If someone close to me had a mental disorder, I would encourage her/him to look for a psychiatrist.”, and
d. self-help strategies (e.g., “Physical exercise contributes to good mental health.”; “Doing something enjoyable contributes to a good mental health.”)

For each item, participants were asked to indicate their level of agreement or disagreement on a five-point Likert scale (ranging from 1—strongly disagree to 5—strongly agree).

A sample of 6,660 participants was assessed across all countries. However, only data from participants aged 17-25 years old, with valid (non-missing) values for gender and for all scale items, was analysed, totalling 5,054.

## Data analysis

A confirmatory factor analysis (CFA) was conducted, stratified by country, to assess the four-factor structure of the 16-item scale. Chi-square (model vs. saturated), Comparative Fit Index (CFI), Root Mean Squared Error of Approximation (RMSEA), Standardized root mean squared residual (SRMR), and Akaike’s information criterion (AIC), were computed to evaluate models’ goodness of fit. A good model fit was defined by RMSEA ≤ .06, SRMR ≤ .08 and CFI ≥ .90^21^.

Standardized factor loadings (standard errors) and the Squared Multiple Correlation Coefficient (R2) retrieved from the stratified models for items were also computed. Internal consistency was analysed using Cronbach’s Alpha. Alpha values between 0.70 and 0.95 was considered good^22^.

Means (standard deviations) and correlations of each MHL dimension and the scales total score were computed for each country. IBM SPSS Statistics v29 was used for descriptives and correlations and Stata v18 was used for CFA.

## Results

Table 1 presents participants’ gender and age by country. The largest sample collected was from Poland (*n* = 1446) and the smallest was from Pakistan (*n* = 144). There were more women participants than men across all countries ranging from 57.0 % in Nepal to 80.1% in Brazil.

**Table 1.** Participants by gender and country included in analysis.

|  | **Men** | **Women** | **Other** | **Total** |
| --- | --- | --- | --- | --- |
|  | n (%) | n (%) | n (%) |  |
| **Greece** | 83 (29.9) | 193 (69.4) | 2 (0.7) | 278 |
| **Poland** | 261 (18.0) | 1104 (76.3) | 81 (5.6) | 1446 |
| **Germany** | 105 (21.2) | 382 (77.2) | 8 (1.6) | 495 |
| **Brazil** | 113 (19.4) | 467 (80.1) | 3 (0.5) | 583 |
| **Turkey** | 174 (30.0) | 406 (70.0) | 0 | 580 |
| **Nigeria** | 48 (27.1) | 128 (72.3) | 1 (0.6) | 177 |
| **Lithuania** | 58 (39.7) | 88 (60.3) | 0 | 146 |
| **Serbia** | 72 (21.0) | 269 (78.4) | 2 (0.6) | 343 |
| **Pakistan** | 59 (41.7) | 85 (58.3) | 0 | 144 |
| **Nepal** | 319 (43.0) | 423 (57.0) | 0 | 742 |
| **Malaysia** | 51 (42.5) | 69 (57.5) | 0 | 120 |
|  | 1343 | 3614 | 97 |  |

As shown in Table 2, the models confirming the scales’ four dimensions showed good indices across all countries. The only exceptions were Brazil, with a higher χ^2^ value (χ2/df=7.533) and a lower CFI (0.798), Pakistan, with a higher Root mean squared error of approximation (RMSEA) and Standardized root mean squared residual (SRMR) (0.134 and 0.162 respectively), and Malaysia, with a higher RMSEA (0.095).

**Table 2.** Model goodness of fit indices for confirmatory factor analysis, by country.

| | $\chi^2$ | $\chi^2 / df$ | CFI | RMSEA (90% CI) | SRMR | AIC |
| --- | --- | --- | --- | --- | --- | --- |
| **Greece** | 221.501 | 2.260 | 0.908 | 0.067 (0.056-0.079) | 0.061 | 10002.591 |
| **Poland** | 363.753 | 3.712 | 0.940 | 0.043 (0.039-0.048) | 0.047 | 51766.613 |
| **Germany** | 215.871 | 2.202 | 0.933 | 0.049 (0.040-0.058) | 0.052 | 16088.399 |
| **Brazil** | 738.264 | 7.533 | 0.798 | 0.106 (0.099-0.113) | 0.080 | 20224.995 |
| **Turkey** | 279.289 | 2.850 | 0.942 | 0.057 (0.049-0.064) | 0.042 | 23822.219 |
| **Nigeria** | 153.823 | 1.570 | 0.896 | 0.057 (0.039-0.074) | 0.070 | 5928.658 |
| **Lithuania** | 165.805 | 1.692 | 0.893 | 0.069 (0.050-0.087) | 0.071 | 5146.150 |
| **Serbia** | 185.490 | 1.893 | 0.932 | 0.051 (0.040-0.062) | 0.058 | 13450.964 |
| **Pakistan** | 349.771 | 3.569 | 0.465 | 0.134 (0.119-0.149) | 0.162 | 5244.484 |
| **Nepal** | 264.224 | 2.696 | 0.899 | 0.048 (0.041-0.055) | 0.042 | 27675.848 |
| **Malaysia** | 202.377 | 2.065 | 0.911 | 0.095 (0.076-0.113) | 0.076 | 3206.994 |

$\chi^2$ – chi-square for model's vs saturated (all were $p < 0.000$ ); $\chi^2 / df$ - Chi-square divided by degrees of freedom; CFI - Comparative fit index; RMSEA- Root mean squared error of approximation; SRMR - Standardized root mean squared residual; AIC - Akaike's information criterion

The factorial weights obtained from the models fitted for CFA (Tables 3a and 3b), were, in general high, although with some nuances for specific items in specific countries. In Greece, Germany, Brazil, Serbia, and Malaysia all standardized factor loadings (SL) were above 0.400. In Poland, item 15 showed a SL of 0.183, but all remaining items were above 0.40. In Turkey, item 27 demonstrated a negative SL (-0.574), whereas in Nigeria and Lithuania, items 1 and 22, showed the lowest SL. In Pakistan, item 7 showed a SL of -0.048 and item 22 a SL of -0.005. In Nepal, items 1, 16, 22 and 27 had lower SLs (ranging from 0.262 to 0.360) whereas the remaining items had SLs higher than 0.418.

**Table 3a.**
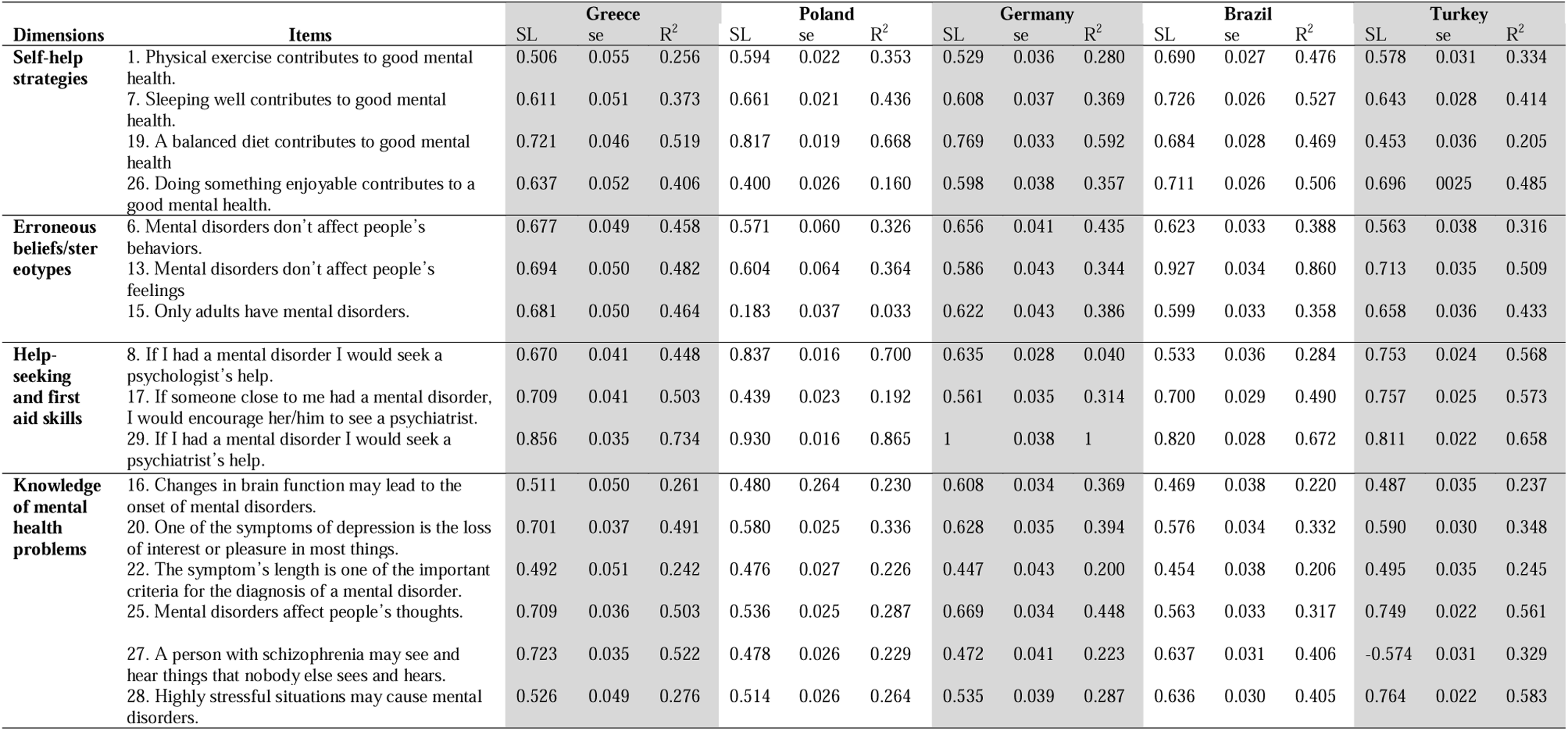
Standardized Factor Loadings (SL), Standardized Error (se) and Squared Multiple Correlations (R^2^) in Greece, Poland, Germany, Brazil and Turkey.

**Table 3b.** Standardized Factor Loadings (SL), Standardized Error (se) and Squared Multiple Correlations (R^2^) in Nigeria, Lithuania, Serbia, Pakistan and Nepal.

| Dimensions | Items | Nigeria |  |  | Lithuania |  |  | Serbia |  |  | Pakistan |  |  | Nepal |  |  |
| --- | --- | --- | --- | --- | --- | --- | --- | --- | --- | --- | --- | --- | --- | --- | --- | --- |
|  |  | SL | se | R<sup>2</sup> | SL | se | R<sup>2</sup> | SL | se | R<sup>2</sup> | SL | se | R<sup>2</sup> | SL | se | R<sup>2</sup> |
| **Self-help strategies** | 1. Physical exercise contributes to good mental health. | 0.349 | 0.102 | 0.122 | 0.386 | 0.087 | 0.149 | 0.627 | 0.043 | 0.393 | 0.417 | 0.091 | 0.174 | 0.262 | 0.041 | 0.068 |
|  | 7. Sleeping well contributes to good mental health. | 0.427 | 0.107 | 0.182 | 0.608 | 0.074 | 0.370 | 0.704 | 0.040 | 0.495 | -0.093 | 0.110 | 0.009 | 0.444 | 0.037 | 0.197 |
|  | 19. A balanced diet contributes to good mental health | 0.559 | 0.109 | 0.312 | 0.722 | 0.068 | 0.522 | 0.660 | 0.010 | 0.435 | 0.618 | 0.082 | 0.381 | 0.547 | 0.034 | 0.300 |
|  | 26. Doing something enjoyable contributes to a good mental health. | 0.652 | 0.094 | 0.425 | 0.551 | 0.080 | 0.304 | 0.616 | 0.045 | 0.379 | 0.573 | 0.085 | 0.328 | 0.639 | 0.033 | 0.408 |
| **Erroneous beliefs/ster eotypes** | 6. Mental disorders don't affect people's behaviors. | 0.494 | 0.096 | 0.244 | 0.552 | 0.073 | 0.305 | 0.489 | 0.065 | 0.240 | 0.581 | 0.078 | 0.338 | 0.547 | 0.047 | 0.299 |
|  | 13. Mental disorders don't affect people's feelings | 0.457 | 0.096 | 0.209 | 0.705 | 0.067 | 0.498 | 0.578 | 0.066 | 0.335 | 0.386 | 0.090 | 0.149 | 0.590 | 0.048 | 0.348 |
|  | 15. Only adults have mental disorders. | 0.521 | 0.095 | 0.271 | 0.793 | 0.066 | 0.629 | 0.578 | 0.067 | 0.309 | 0.652 | 0.078 | 0.425 | 0.466 | 0.046 | 0.217 |
| **Help-seeking and first aid skills** | 8. If I had a mental disorder I would seek a psychologist's help. | 0.722 | 0.045 | 0.521 | 0.772 | 0.044 | 0.595 | 0.832 | 0.027 | 0.693 | 0.528 | 0.077 | 0.279 | 0.590 | 0.032 | 0.348 |
|  | 17. If someone close to me had a mental disorder, I would encourage her/him to see a psychiatrist. | 0.671 | 0.050 | 0.450 | 0.690 | 0.052 | 0.476 | 0.560 | 0.040 | 0.313 | 0.502 | 0.081 | 0.252 | 0.684 | 0.030 | 0.467 |
|  | 29. If I had a mental disorder I would seek a psychiatrist's help. | 0.935 | 0.039 | 0.874 | 0.928 | 0.038 | 0.861 | 0.978 | 0.025 | 0.956 | 0.345 | 0.091 | 0.119 | 0.689 | 0.030 | 0.474 |
| **Knowledge of mental health problems** | 16. Changes in brain function may lead to the onset of mental disorders. | 0.415 | 0.074 | 0.172 | 0.593 | 0.069 | 0.352 | 0.483 | 0.053 | 0.233 | 0.851 | 0.122 | 0.724 | 0.337 | 0.040 | 0.113 |
|  | 20. One of the symptoms of depression is the loss of interest or pleasure in most things. | 0.615 | 0.062 | 0.378 | 0.638 | 0.062 | 0.407 | 0.526 | 0.051 | 0.277 | 0.251 | 0.101 | 0.063 | 0.418 | 0.039 | 0.175 |
|  | 22. The symptom's length is one of the important criteria for the diagnosis of a mental disorder. | 0.359 | 0.077 | 0.129 | 0.406 | 0.082 | 0.165 | 0.425 | 0.055 | 0.181 | 0.608 | 0.084 | 0.370 | 0.360 | 0.040 | 0.130 |
|  | 25. Mental disorders affect people's thoughts. | 0.599 | 0.063 | 0.359 | 0.690 | 0.059 | 0.477 | 0.585 | 0.048 | 0.342 | 0.158 | 0.117 | 0.025 | 0.458 | 0.037 | 0.210 |
|  | 27. A person with schizophrenia may see and hear things that nobody else sees and hears. | 0.604 | 0.063 | 0.365 | 0.608 | 0.065 | 0.369 | 0.594 | 0.048 | 0.352 | 0.138 | 0.099 | 0.019 | 0.333 | 0.041 | 0.111 |
|  | 28. Highly stressful situations may cause mental disorders. | 0.404 | 0.075 | 0.163 | 0.518 | 0.072 | 0.268 | 0.538 | 0.050 | 0.289 | 0.041 | 0.118 | 0.002 | 0.569 | 0.034 | 0.323 |

| Dimensions | Items | Malaysia |  |  |
| --- | --- | --- | --- | --- |
|  |  | SL | se | R<sup>2</sup> |
| **Self-help strategies** | 1. Physical exercise contributes to good mental health. | 0.785 | 0.041 | 0.626 |
|  | 7. Sleeping well contributes to good mental health. | 0.902 | 0.025 | 0.814 |
|  | 19. A balanced diet contributes to good mental health | 0.850 | 0.032 | 0.722 |
|  | 26. Doing something enjoyable contributes to a good mental health. | 0.856 | 0.030 | 0.733 |
| **Erroneous beliefs/stereotypes** | 6. Mental disorders don't affect people's behaviors. | 0.910 | 0.046 | 0.894 |
|  | 13. Mental disorders don't affect people's feelings | 0.828 | 0.049 | 0.686 |
|  | 15. Only adults have mental disorders. | 0.640 | 0.062 | 0.409 |
| **Help-seeking and first aid skills** | 8. If I had a mental disorder I would seek a psychologist's help. | 0.834 | 0.038 | 0.695 |
|  | 17. If someone close to me had a mental disorder, I would encourage her/him to see a psychiatrist. | 0.664 | 0.059 | 0.441 |
|  | 29. If I had a mental disorder I would seek a psychiatrist's help. | 0.909 | 0.034 | 0.827 |
| **Knowledge of mental health problems** | 16. Changes in brain function may lead to the onset of mental disorders. | 0.716 | 0.051 | 0.512 |
|  | 20. One of the symptoms of depression is the loss of interest or pleasure in most things. | 0.868 | 0.031 | 0.753 |
|  | 22. The symptom's length is one of the important criteria for the diagnosis of a mental disorder. | 0.731 | 0.049 | 0.534 |
|  | 25. Mental disorders affect people's thoughts. | 0.780 | 0.042 | 0.608 |
|  | 27. A person with schizophrenia may see and hear things that nobody else sees and hears. | 0.701 | 0.052 | 0.491 |
|  | 28. Highly stressful situations may cause mental disorders. | 0.784 | 0.041 | 0.615 |

Cronbach’s alpha computed to estimate scale’s reliability is shown in Table 4, for the total scale and each dimension, by country. All countries showed acceptable alpha coefficient values for the total scale above 0.64, except Pakistan which had the lowest values (alpha: 0.49).

**Table 4.**
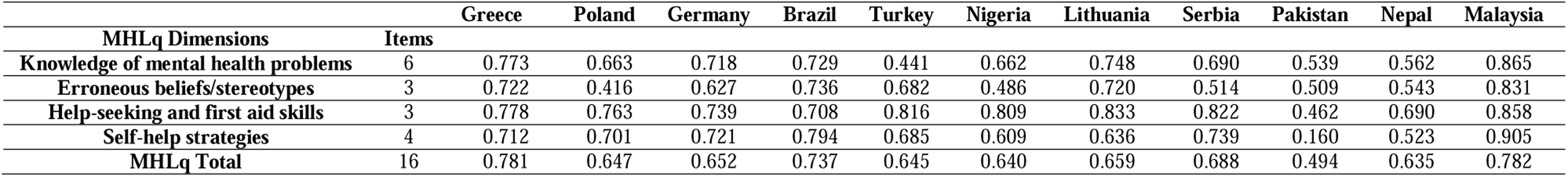
Cronbach’s alpha coefficients for the Mental Health Literacy questionnaire (MHLq) young adults-Short Version.

Table 5 displays the means with standard deviations, and correlations among the scales’ dimensions and the total score by country. Negative correlations were observed between the dimension “Erroneous beliefs/stereotypes” and the other dimensions and total score, reflecting the expected pattern of answers for the set of items included in this dimension. The correlations between the dimensions “Knowledge of mental health problems”, “Help-seeking and first aid skills”, “Self-help strategies” and the total score were strong (>0.64) and statistically significant (*p* < 0.001) across all countries. However, the correlation of the erroneous belief/stereotypes subscale with the MHLq total score was significantly lower in all countries (-0.180 to 0.166) compared to the other subscales (0.616 to 0.846).

**Table 5.** Means, Standard Deviations (sd) and Correlations Among the Mental Health Literacy questionnaire (MHLq) Dimensions and Total Score in Greece, Poland, Germany, Brazil, Turkey, Nigeria, Lithuania, Serbia, Pakistan and Nepal.

|  |  | Greece |  |  |  |
| --- | --- | --- | --- | --- | --- |
|  | Mean (sd) | F2 | F3 | F4 | Total |
| F1 - Knowledge of mental health problems | 4.009 (0.555) | -0.247** | 0.518** | 0.619** | 0.825** |
| F2 - Erroneous beliefs/stereotypes | 1.628 (0.735) |  | -0.063 | -0.105 | 0.150 |
| F3 - Help-seeking and first aid skills | 4.169 (0.780) |  |  | 0.389** | 0.733** |
| F4 - Self-help strategies | 4.172 (0.589) |  |  |  | 0.772** |
| MHLq Total | 3.633 (0.413) |  |  |  |  |
|  |  | Poland |  |  |  |
|  | Mean (sd) | F2 | F3 | F4 | Total |
| F1 - Knowledge of mental health problems | 4.468 (0.442) | -0.085** | 0.285** | 0.274** | 0.679** |
| F2 - Erroneous beliefs/stereotypes | 1.480 (0.740) |  | -0.066* | -0.060* | 0.193** |
| F3 - Help-seeking and first aid skills | 4.090 (0.838) |  |  | 0.214** | 0.643** |
| F4 - Self-help strategies | 4.035 (0.636) |  |  |  | 0.644** |
| MHLq Total | 3.729 (0.357) |  |  |  |  |
|  |  | Germany |  |  |  |
|  | Mean (sd) | F2 | F3 | F4 | Total |
| F1 - Knowledge of mental health problems | 4.304 (0.482) | -0.328** | 0.216** | 0.373** | 0.734** |
| F2 - Erroneous beliefs/stereotypes | 1.339 (0.534) |  | -0.131** | -0.179** | -0.003 |
| F3 - Help-seeking and first aid skills | 4.123 (0.772) |  |  | 0.196** | 0.634** |
| F4 - Self-help strategies | 4.323 (0.508) |  |  |  | 0.664** |
| MHLq Total | 3.719 (0.309) |  |  |  |  |
|  |  | Brazil |  |  |  |
|  | Mean (sd) | F2 | F3 | F4 | Total |
| F1 - Knowledge of mental health problems | 4.204 (0.529) | -0.191** | 0.529** | 0.566** | 0.857** |
| F2 - Erroneous beliefs/stereotypes | 1.512 (0.771) |  | -0.216** | -0.332** | 0.083* |
| F3 - Help-seeking and first aid skills | 4.274 (0.682) |  |  | 0.420** | 0.702** |
| F4 - Self-help strategies | 4.445 (0.579) |  |  |  | 0.703** |
| MHLq Total | 3.770 (0.374) |  |  |  |  |
|  |  | Turkey |  |  |  |
|  | Mean (sd) | F2 | F3 | F4 | Total |
| F1 - Knowledge of mental health problems | 3.645 (0.509) | -0.368** | 0.495** | 0.651** | 0.807** |
| F2 - Erroneous beliefs/stereotypes | 1.715 (0.870) |  | -0.366** | -0.401** | -0.095* |
| F3 - Help-seeking and first aid skills | 4.094 (0.890) |  |  | 0.579** | 0.747** |
| F4 - Self-help strategies | 4.118 (0.704) |  |  |  | 0.816** |
| MHLq Total | 3.968 (0.545) |  |  |  |  |
|  |  | Nigeria |  |  |  |
|  | Mean (sd) | F2 | F3 | F4 | Total |
| F1 - Knowledge of mental health problems | 4.029 (0.437) | -0.286** | 0.346** | 0.444** | 0.761** |
| F2 - Erroneous beliefs/stereotypes | 1.721 (0.607) |  | -0.099 | -0.178* | 0.115 |
| F3 - Help-seeking and first aid skills | 4.058 (0.645) |  |  | 0.232** | 0.658** |
| F4 - Self-help strategies | 4.130 (0.469) |  |  |  | 0.668** |
| MHLq Total | 3.627 (0.297) |  |  |  |  |
|  |  | Lithuania |  |  |  |
|  | Mean (sd) | F2 | F3 | F4 | Total |
| F1 - Knowledge of mental health problems | 3.950 (0.478) | -0.341** | 0.262** | 0.449** | 0.712** |
| F2 - Erroneous beliefs/stereotypes | 1.772 (0.699) |  | -0.088 | -0.282** | 0.052 |
| F3 - Help-seeking and first aid skills | 3.731 (0.898) |  |  | 0.206* | 0.690** |
| F4 - Self-help strategies | 4.062 (0.537) |  |  |  | 0.633** |
| MHLq Total | 3.528 (0.336) |  |  |  |  |
| Serbia |  |  |  |  |  |
|  | Mean (sd) | F2 | F3 | F4 | Total |
| F1 - Knowledge of mental health problems | 4.171 (0.562) | -0.271** | 0.162* | 0.377** | 0.686** |
| F2 - Erroneous beliefs/stereotypes | 1.428 (0.623) |  | -0.020 | -0.080 | 0.107* |
| F3 - Help-seeking and first aid skills | 4.166 (0.993) |  |  | 0.234** | 0.646** |
| F4 - Self-help strategies | 4.197 (0.685) |  |  |  | 0.714** |
| MHLq Total | 3.661 (0.399) |  |  |  |  |
| Pakistan |  |  |  |  |  |
|  | Mean (sd) | F2 | F3 | F4 | Total |
| F1 - Knowledge of mental health problems | 3.551 (0.485) | -0.340** | 0.424** | 0.422** | 0.846** |
| F2 - Erroneous beliefs/stereotypes | 1.692 (0.496) |  | -0.439** | -0.342** | -0.180* |
| F3 - Help-seeking and first aid skills | 4.261 (0.463) |  |  | 0.408** | 0.616** |
| F4 - Self-help strategies | 3.769 (0.519) |  |  |  | 0.735** |
| MHLq Total | 3.390 (0.286) |  |  |  |  |
| Nepal |  |  |  |  |  |
|  | Mean (sd) | F2 | F3 | F4 | Total |
| F1 - Knowledge of mental health problems | 3.904 (0.474) | -0.159** | 0.421** | 0.407** | 0.784** |
| F2 - Erroneous beliefs/stereotypes | 1.752 (0.659) |  | -0.189** | -0.150** | 0.166** |
| F3 - Help-seeking and first aid skills | 4.356 (0.578) |  |  | 0.457** | 0.671** |
| F4 - Self-help strategies | 4.297 (0.540) |  |  |  | 0.722** |
| MHLq Total | 3.684 (0.330) |  |  |  |  |
| Malaysia |  |  |  |  |  |
|  | Mean (sd) | F2 | F3 | F4 | Total |
| F1 - Knowledge of mental health problems | 4.330 (0.510) | -0.355** | 0.444** | 0.508** | 0.735** |
| F2 - Erroneous beliefs/stereotypes | 1.890 (0.834) |  | -0.172** | -0.182** | 0.115* |
| F3 - Help-seeking and first aid skills | 4.325 (0.664) |  |  | 0.528** | 0.719** |
| F4 - Self-help strategies | 4.492 (0.567) |  |  |  | 0.772** |
| MHLq Total | 3.912 (0.358) |  |  |  |  |

\* $p < .05$ ; \*\* $p < .01$ ; Items composing F2 – Erroneous beliefs/stereotypes, were not inverted in this analysis (for this subscale, lower scores indicate less agreement with Erroneous beliefs/stereotypes).

## Discussion

This study examined the psychometric properties of the 16-item short version of the Mental Health Literacy questionnaire for young adults (MHLq-SVa) across samples from Greece, Poland, Germany, Brazil, Turkey, Nigeria, Lithuania, Serbia, Pakistan, Malaysia, and Nepal. By including diverse populations from four continents, this research expands the multicultural validity of the MHLq-SVa, extending its psychometric evaluation to a broad range of languages and cultural contexts.

The results suggest that the four-dimensional solution of the scale has good psychometric properties across all countries, including acceptable internal consistency according to established reference values^22^. However, some goodness-of-fit indices suggested poorer performance in Pakistan, which was confirmed by low values noted for the standardized loadings of specific items. A recent systematic review on MHL in Pakistan suggested the lack of prevalence studies and comparability of MHL measures and reflected on how different cultural beliefs associated with mental health (e.g. preference for religious healers, naturopathy practitioners, homeopathic doctors, or faith healers who treat mental health problems) could impact MHL ^23^. Further studies should explore how such factors influence cross-cultural differences in MHL reports. Nevertheless, the mean scores obtained in Pakistan for the four scales’ dimensions and their interrelations were well aligned with the results obtained in all other countries analysed, suggesting consistency of the scale in this sample.

Slightly poorer performance as suggested by some of the goodness-of-fit indices was observed in Brazil. To this global goodness-of-fit indices could have influenced the data collection method, which was uncontrolled by the researchers. However, when looking at the Brazilian individual standardized items loadings, values ranged between 0.454 and 0.972, suggesting overall congruency and similarity with loadings obtained in the other countries.

Some limitations of the current study should be mentioned. The methods of administration were diverse (online, paper-and-pencil), which can result, for example, in some selection bias, if only people with access to the questionnaire online could participate in any specific country. In each site, researchers added different questions before or after the MHLq scale, according to their interests for further explorations. Because of such additions (from few items to other full scales fitting national-level goals), the positioning of the MHLq within longer interviews may have impacted the burden associated with its completion, leading to fatigue or priming effects, or to socially desirable responses towards the end of the questionnaire.

The differences in recruitment, sampling and sample sizes by country could have impacted these findings’ external validity and statistical power, respectively. Future research should, ideally, use probability sampling procedures aiming for representativeness, assess larger samples, and compare, whenever possible, the results obtained through different modes of administration (preferably within the same country or sample).

The fact that most data was collected during or after the COVID-19 pandemic could have impacted the results, since awareness of mental health-related problems generally increased during this period, following numerous studies, surveys, campaigns and support enacted^24,25^. This calls for repeated assessments of MHL levels using the same scale to more clearly disentangle the influence of such recent surge in mental health and pandemic-related information worldwide.

Future studies should also explore the scales’ concurrent or criterion validity by applying other MHL standardized measures and exploring its results in demographically mixed samples ^22^.

To effectively promote mental health and mitigate the burden of mental ill health it is paramount to increase MHL levels of the population. This plays a pivotal role in early identification and decision to seek appropriate treatment and/or support. The Mental Health Literacy questionnaire – short version, as a standardized tool to measure this construct, can be an important ally for researchers, practitioners, or other public mental health stakeholders, by maximizing cross-cultural comparability and allowing the monitoring of MHL key dimensions following relevant interventions.

## Data Availability

The datasets used and/or analysed during the current study are available from the corresponding author on reasonable request and following consultation and agreement with the group of authors from each country that contributed to this manuscript.

## Acknowledgments

The authors thank the participants for actively participating in the study.

## Statements of ethical approval

All participants provided online consent, except in Turkey, where participants provided written consent. Furthermore, ethical standards of the institutional and/or national research committees and with the 1964 Helsinki Declaration and its later amendments or comparable ethical standards, were followed in all countries.

Ethical approval was obtained from local ethics committees in each country, except in Nepal and Poland where this was not a requirement. In Brazil, the study protocol was reviewed by the Research Ethics Committee of the University of São Paulo (reference: 3.995.379), in Germany by the Ethics Commission of JGU Mainz and the State Chamber of Physicians (refs: 2022-JGU-psychEK-S041; 2023-JGU-psychEK-S017; 2023-JGU-psychEK-005 and 2022-16888-other), in Greece, by the Ethics committee of the Hellenic Mediterranean University (ref: 42(130)/25-10-2022), in Lithuania (whose corresponding researcher coordinated data collection in Lithuania and Nigeria) by the Ethics Committee of Klaipeda University (Faculty of Health Science, Public Health Department), In Malaysia by the Medical Research & Ethics Committee (ref: NMRR ID-22-01887-XCU (IIR)), in Pakistan by the Committee for Evaluation of Research Projects, University of the Punjab, Lahore, Pakistan (No. D/ 503 /Est- I dated: 14/02/2020), in Serbia by the Committee for Assessment of Ethicality in Scientific Research of the Institute for Educational Research (ref: 163/2023), and in Turkey by the Ordu University Ethics Committee (ref: KAEK-58-2019-68).

## Authors’ contributions

LC and PD originated the idea. DC performed data analysis. All authors, except DC, contributed to data collection. DC, MC, LC, and PD wrote the first manuscript draft. All authors participated in the critical review of the manuscript and read and approved the final manuscript.

## References

1. Global, regional, and national burden of 12 mental disorders in 204 countries and territories, 1990–2019: a systematic analysis for the Global Burden of Disease Study 2019. Lancet Psychiatry. 2022 Feb;9(2):137–50.

2. Wu Y, Wang L, Tao M, Cao H, Yuan H, Ye M, et al. Changing trends in the global burden of mental disorders from 1990 to 2019 and predicted levels in 25 years. Epidemiol Psychiatr Sci. 2023 Nov 7;32:e63.

3. Henderson C, Evans-Lacko S, Thornicroft G. Mental Illness Stigma, Help Seeking, and Public Health Programs. Am J Public Health. 2013 May;103(5):777–80.

4. Rüsch N, Corrigan PW, Wassel A, Michaels P, Larson JE, Olschewski M, et al. Self-stigma, group identification, perceived legitimacy of discrimination and mental health service use. British Journal of Psychiatry. 2009 Dec 2;195(6):551–2.

5. Fekih-Romdhane F, Jahrami H, Stambouli M, Alhuwailah A, Helmy M, Shuwiekh HAM, et al. Cross-cultural comparison of mental illness stigma and help-seeking attitudes: a multinational population-based study from 16 Arab countries and 10,036 individuals. Soc Psychiatry Psychiatr Epidemiol. 2023 Apr 30;58(4):641–56.

6. Torales J, Aveiro-Róbalo TR, Ríos-González C, Barrios I, Almirón-Santacruz J, González-Urbieta I, et al. Discrimination, stigma and mental health: what’s next? International Review of Psychiatry. 2023 May 19;35(3–4):242–50.

7. Evans-Lacko S, Brohan E, Mojtabai R, Thornicroft G. Association between public views of mental illness and self-stigma among individuals with mental illness in 14 European countries. Psychol Med. 2012 Aug 16;42(8):1741–52.

8. Jorm AF. Mental health literacy: Empowering the community to take action for better mental health. American Psychologist. 2012 Apr;67(3):231–43.

9. Cotton SM, Wright A, Harris MG, Jorm AF, Mcgorry PD. Influence of Gender on Mental Health Literacy in Young Australians. Australian & New Zealand Journal of Psychiatry. 2006 Sep 1;40(9):790–6.

10. Reavley NJ, McCann T V., Jorm AF. Mental health literacy in higher education students. Early Interv Psychiatry. 2012 Feb 20;6(1):45–52.

11. Swami V. Mental Health Literacy of Depression: Gender Differences and Attitudinal Antecedents in a Representative British Sample. PLoS One. 2012 Nov 14;7(11):e49779.

12. Doll CM, Michel C, Betz LT, Schimmelmann BG, Schultze-Lutter F. The Important Role of Stereotypes in the relation between Mental Health Literacy and Stigmatization of Depression and Psychosis in the Community. Community Ment Health J. 2022 Apr 26;58(3):474–86.

13. Iswanto ED, Ayubi D. The Relationship of Mental Health Literacy to Help-Seeking Behavior: Systematic Review. Journal of Social Research. 2023 Feb 10;2(3):755–64.

14. Yap MBH, Reavley NJ, Jorm AF. Associations between stigma and help-seeking intentions and beliefs: Findings from an Australian national survey of young people. Psychiatry Res. 2013 Dec;210(3):1154–60.

15. Furnham A, Hamid A. Mental health literacy in non-western countries: a review of the recent literature. Mental Health Review Journal. 2014 Jun 3;19(2):84–98.

16. Wang C, Li H, Li L, Xu D, Kane RL, Meng Q. Health literacy and ethnic disparities in health-related quality of life among rural women: results from a Chinese poor minority area. Health Qual Life Outcomes. 2013 Dec 11;11(1):153.

17. ElKhalil R, AlMekkawi M, O’Connor M, Masuadi E, Sherif M, Belfakir M, et al. Measurement properties of the Mental Health Literacy Scale (MHLS): A systematic review. Asian J Psychiatr [Internet]. 2024 Nov;101:104214. Available from: https://linkinghub.elsevier.com/retrieve/pii/S1876201824003071

18. Dias P, Campos L, Almeida H, Palha F. Mental Health Literacy in Young Adults: Adaptation and Psychometric Properties of the Mental Health Literacy Questionnaire. Int J Environ Res Public Health. 2018 Jun 23;15(7):1318.

19. Campos L, Dias P, Costa M, Rabin L, Miles R, Lestari S, et al. Mental health literacy questionnaire-short version for adults (MHLq-SVa): validation study in China, India, Indonesia, Portugal, Thailand, and the United States. BMC Psychiatry. 2022 Nov 16;22(1):713.

20. Erkut S. Developing Multiple Language Versions of Instruments for Intercultural Research. Child Dev Perspect. 2010 Apr 11;4(1):19–24.

21. Hu L, Bentler PM. Cutoff criteria for fit indexes in covariance structure analysis: Conventional criteria versus new alternatives. Struct Equ Modeling. 1999 Jan;6(1):1– 55.

22. Terwee CB, Bot SDM, de Boer MR, van der Windt DAWM, Knol DL, Dekker J, et al. Quality criteria were proposed for measurement properties of health status questionnaires. J Clin Epidemiol. 2007 Jan;60(1):34–42.

23. Munawar K, Abdul Khaiyom JH, Bokharey IZ, Park MS, Choudhry FR. A systematic review of mental health literacy in Pakistan. Asia-Pacific Psychiatry. 2020 Dec 17;12(4).

24. Else H. How a torrent of COVID science changed research publishing — in seven charts. Nature. 2020 Dec 24;588(7839):553–553.

25. The Lancet Psychiatry. COVID-19 and mental health. Lancet Psychiatry [Internet]. 2021 Feb;8(2):87. Available from: https://linkinghub.elsevier.com/retrieve/pii/S2215036621000055

